# Search for a genetic cause of variably protease-sensitive prionopathy

**DOI:** 10.1101/2024.12.12.24318867

**Authors:** Yuan Lian, Keisi Kotobelli, Stacey Hall, Michael E Talkowski, Anne O’Donnell-Luria, Sonia M Vallabh, Brian S Appleby, Eric Vallabh Minikel

## Abstract

Variably protease-sensitive prionopathy (VPSPr) is a rare, atypical subtype of prion disease currently classified as sporadic. We performed exome sequencing and targeted sequencing of *PRNP* non-coding regions on genomic DNA from autopsy-confirmed VPSPr patients (N=67) in order to search for a possible genetic cause. Our search identified no potentially causal variants for VPSPr. The common polymorphism *PRNP* M129V was the largest genetic risk factor for VPSPr, with an odds ratio of 7.0. Other variants in and near *PRNP* exhibited association to VPSPr risk only in proportion to their linkage disequilibrium with M129V, and upstream expression quantitative trait loci showed no evidence of independent association to VPSPr risk. We cannot rule out the possibility of causal variants hiding in genomic regions or classes of genetic variation that our search did not canvas. Nevertheless, our data support the classification of VPSPr as a sporadic prion disease.

**Author Summary:** Prion disease is caused by misfolding of the prion protein (PrP), and can be either sporadic genetic, or acquired. Acquired cases arising from infection through dietary or medical routes are exceedingly rare today (<1% of cases). Sporadic cases occur apparently at random, without any major genetic risk factors, and are not passed down to subsequent generations. All cases of genetic prion disease to date have been traced to DNA changes that alter the amino acid sequence of PrP, and children of people with genetic prion disease are at 50/50 risk of inheriting these autosomal dominant DNA changes. Variably protease sensitive prionopathy, or VPSPr, is a rare and unusual subtype of prion disease, classified as sporadic because there are no changes in PrP’s amino acid sequence. Here we performed DNA sequencing on 67 VPSPr cases to determine whether a genetic variant outside of the PrP amino acid sequence might cause the disease. We found no DNA changes that could potentially cause VPSPr. While it is difficult to prove the negative — a causal genetic change could still exist in some part of the genome where we did not search — our data support the notion that VPSPr is truly sporadic in nature, and that risk of VPSPr is not transmitted in families.

## Introduction

Prion disease is an invariably fatal neurodegenerative disease caused by autocatalytic templating of the prion protein (PrP) into a misfolded conformer known as a prion [1]. Early on, the generation of PrP species resistant to limited protease digestion was considered an obligate pathological hallmark of prion disease [2–4]. Later, it was determined that only some proportion of misfolded pathologic PrP is protease-resistant, and that this proportion differs between prion strains [5]. Variably protease-sensitive prionopathy (VPSPr) is a subtype of human prion disease named for the paucity or total absence of protease-resistant PrP in the brain [6,7]. VPSPr possesses several additional features that distinguish it from other types of human prion disease [8]. Histopathologically, it features larger spongiform vacuoles than sporadic Creutzfeldt-Jakob disease (sCJD), and a distinct pattern of PrP deposition. Transmission to humanized mice has been challenging [9,10], although the transmissibility of VPSPr has been established in bank voles [11]. Cerebrospinal fluid (CSF) real-time quaking induced conversion (RT-QuIC) is less sensitive than in sCJD, being positive in 7 out of 10 cases in the literature [12–15], and sometimes only with low peak amplitude. VPSPr progresses slower than sCJD, with a disease course averaging 2.5 years [7], and is often misdiagnosed as a non-Alzheimer’s disease dementia. Many VPSPr patients exhibit co-occurring tau, amyloid beta, or synuclein pathology [8,16–20].

Most prion disease cases (85%) are sporadic, a term which in prion disease is defined to mean lacking any known environmental or molecular genetic cause. A minority (15%) are genetic, caused by *PRNP* protein-coding variants, some but not all of which are highly penetrant [21]. A smaller minority of cases (<1%) are acquired through infection, via exposure to misfolded prions in the diet or through medical procedures [22]. *PRNP* M129V (rs1799990), a common polymorphism, modifies both disease risk and phenotypic features across all three etiologies [23]. In sCJD, the 129M allele and particularly the 129MM genotype are overrepresented, whereas in VPSPr, the 129V allele is overrepresented. Protein-coding variants in *PRNP* have been ruled out in all reported VPSPr patients, leading to classification of VPSPr as a sporadic, rather than genetic, prion disease. Nevertheless, a large proportion of VPSPr patients have family history positive for dementia — 15/36 (42%) in the largest reported cohort [8], including a majority (60%) of cases with the 129VV genotype. Although controlled studies have not been done to determine whether this rate of family history of dementia differs from other diagnoses, it is nominally higher than the 12.1% reported in sporadic CJD [24], leading to speculation that VPSPr might be a form of genetic prion disease with a cause other than *PRNP* coding variants [7,8].

In this study, we set out to identify a genetic cause of VPSPr. We evaluated the potential genetic architecture of VPSPr by examining disease prevalence and the rate at which positive family history is observed. We determined that if a causal genetic variant exists, it might be only modestly penetrant, yet extremely rare and conferring a high fold increase in lifetime risk of prion disease, with odds ratio (OR) >1,000. Such a variant could be discovered through sequencing tens of cases and comparing their allele frequencies to those of publicly available population control datasets. We obtained DNA from 67 autopsy-confirmed VPSPr cases and performed exome sequencing and deep sequencing of 152 kb centered on the *PRNP* locus, including non-coding regions, and compared these to the Genome Aggregation Database (gnomAD) and controls. No potentially causal variants were identified, suggesting that VPSPr may be genuinely sporadic in etiology.

## Results

### Hypothesized genetic architecture of VPSPr and rationale for study design

To define a search strategy for genomic variants that could hypothetically cause VPSPr, we first needed to consider how rare the causal genetic variants would need to be, and how strongly enriched in cases over controls they would need to be (Table 1). While many genetic architectures for VPSPr are possible, as a starting point, we considered the possibility that the 42% rate of positive family history is due to a modestly penetrant single genetic variant. In the simplified case of 1 variant-positive relative, the family history rate could be explained by a single variant with 42% penetrance. The total prevalence of all forms of prion disease is estimated at 1 in 6,239 deaths, or 0.016% lifetime risk [25]. VPSPr in turn has comprised 88/3,931 prion disease cases (2.2%) ascertained by the U.S. National Prion Disease Pathology and Surveillance Center since the discovery of VPSPr [26]. The highest allele frequency (AF) of a causal variant would occur if 100% of VPSPr cases were caused by a single variant.

**Table 1.** Parameters used to calculate the maximum credible allele frequency for VPSPr.

| Parameter | Estimate | Rationale |
| --- | --- | --- |
| Penetrance | 42% | Most conservative assumption, assuming just 1 variant-positive relative who has lived through the risk period |
| Prevalence of all prion disease | 0.016% | 1 in 6,239 deaths [25] |
| VPSPr as proportion of prion disease | 2.2% | 88/3,931 cases in U.S. since discovery of VPSPr in 2008 [26] |
| Proportion of cases attributable to most common variant | 100% | Most conservative assumption because it yields the highest tolerated allele frequency |
| Maximum credible allele frequency | 4.3e-6 | Calculated from above parameters using the approach of Whiffin & Minikel et al [27] |

We included all of these assumptions (Table 1) into a previously described framework [27] for determining the highest allele frequency that a disease-causing variant could have in the general population, and obtained a maximum credible AF of 4.3e-6. Note that allowing multiple causal variants, no single one of which explains all cases, would only further lower this frequency. Considering multiplex families, where a 42% family history rate could arise from, for example, 2 variant-positive relatives with penetrance 21%, would also lower this frequency. Moreover, our filtering strategy is inherently conservative because it only removes genetic variants when even the lower bound of the 95% confidence interval on their allele frequency is above the filtering threshold. Based on these considerations, we selected 4.3e-6, or 1 in 232,558, as our AF filtering threshold. This seems plausible because VPSPr is several times rarer than genetic prion disease caused by highly penetrant *PRNP* coding variants, which collectively are estimated to have a genetic prevalence of ~1 in 50,000 people [28,29].

We explored the relationship between positive family history and OR for known causal *PRNP* variants in genetic prion disease (Figure 1A). Updating our OR estimates using gnomAD v4.1 data [30] and comparing to the proportion of cases with family history using previously reported data [21], we fit a log-linear model. VPSPr, if its 42% positive family history were indeed explained by a genetic cause, could plausibly arise from variants with OR > 1,000. The constraints of OR > 1,000 and AF < 4.3e-6 define a range of possible positions that a VPSPr-causing variant could occupy in the scatterplot of AF in the general population versus AF in VPSPr cases (Figure 1B). Assuming this hypothesized genetic architecture, we determined that sequencing VPSPr cases alone would be a valid strategy to identify causal variants. A genome-wide association study (GWAS), which is designed to identify common variants (AF > 0.1%) of small effect (OR < 2) would not be appropriate. A case/control exome study, which may identify slightly rarer variants (AF > 0.01%) with slightly larger effects (OR 2-10) is also not appropriate. Instead, the same strategy used for solving highly penetrant Mendelian diseases — sequencing only cases and filtering their genetic variants based on population AF information — is appropriate.

**Figure 1.**
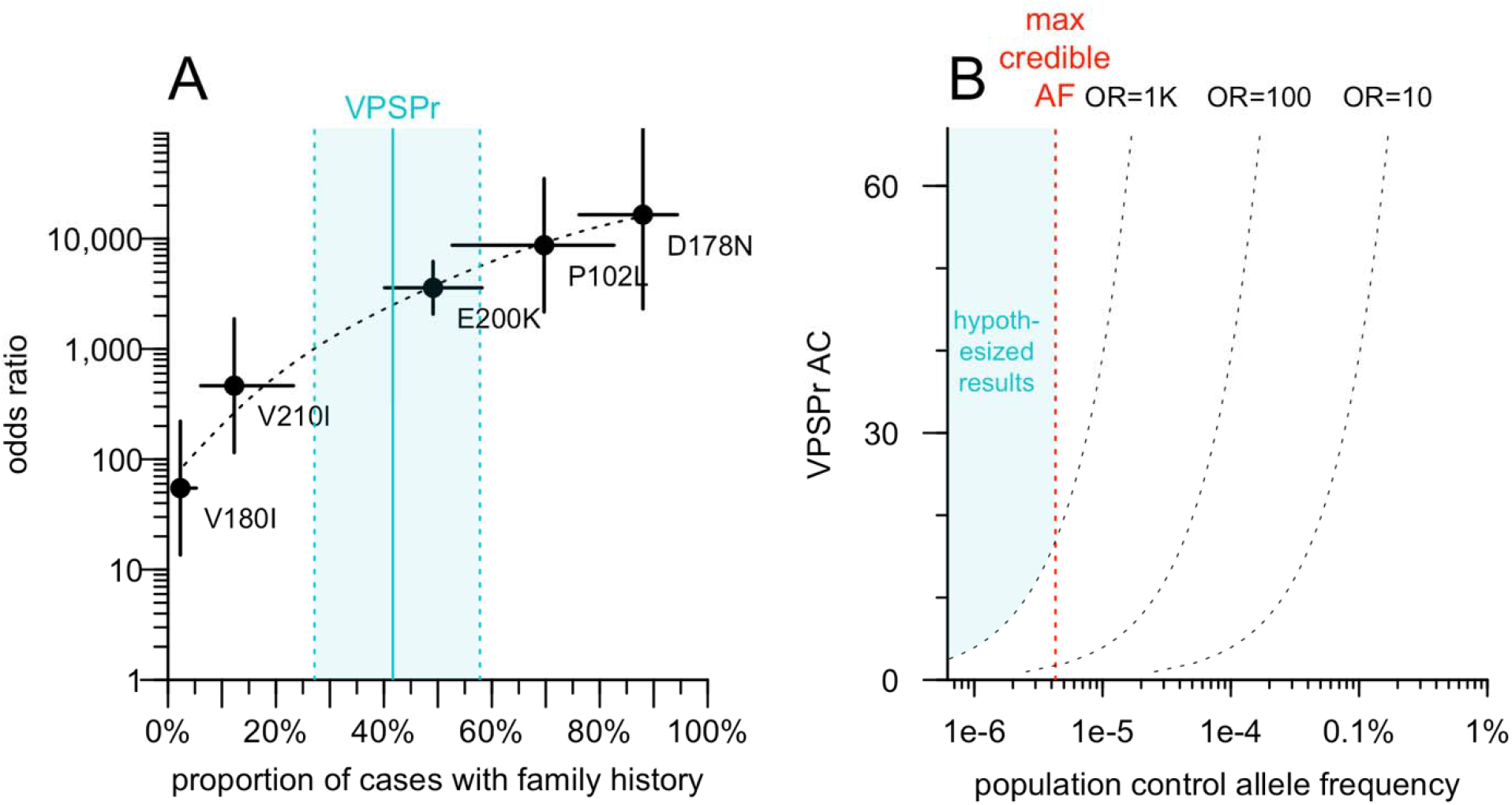
Hypothetical odds ratio of VPSPr-causing variants and implications for genomic search strategy. **A)** Odds ratio (OR; Fisher exact test) for PRNP coding variants that cause genetic prion disease, based on previously reported case data [21] and gnomAD v4.1 allele frequencies, versus prevalence of family history among cases as reported [21]. **B)** Range of possible outcomes for VPSPr-causing variants, see text for reasoning.

### Characteristics of VPSPr cohort

We obtained DNA from N=67 autopsy-confirmed VPSPr cases from the U.S. National Prion Disease Pathology Surveillance Center (NPDPSC; Table 2). All 3 *PRNP* codon 129 genotypes were represented in our cohort, with 129VV comprising a majority (67%, 45/67) of cases, consistent with prior reports. Age at death differed between genotypes (P = 0.0012, Type I ANOVA, Table 2), shifting younger as the number of V alleles increased (Tukey post-hoc test: P = 0.0031 for VV vs. MM, P = 0.051 for VV vs. MV, P = 0.54 for MV vs. MM). Disease duration was also modified by codon 129 genotype, being longest for heterozygous MV individuals (P = 5.0e-6, Type I ANOVA, Table 2), similar to other forms of prion disease [23] (Tukey post-hoc test: P =4.7e-6 for VV vs. MV, P = 0.11 for VV vs. MM, P = 0.18 for MV vs. MM). The sex distribution did not deviate significantly from 50/50 (P = 0.46, binomial test), and sex was not associated to age at death (P = 0.42, Type I ANOVA) nor disease duration (P = 0.57, Type I ANOVA) and showed no confounding with codon 129 genotype (P = 0.28, Fisher exact test). The reporting physician indicated the presence or absence of family history of neurological disease in 19 cases, of whom 10 (52%) had a positive family history, all of whom were 129VV.

**Table 2.** Characteristics of VPSPr DNA cohort.

| **Codon 129 genotype** | **N** | **Age at death (years, mean±SD)** | **Disease duration (years, mean±SD)** | **Sex** | **Family history of neurological disease** |
| --- | --- | --- | --- | --- | --- |
| MM | 9 | 79±8 | 2.7±0.7 | 7M/2F | 0% (0/1) |
| MV | 13 | 75±10 | 3.8±1.7 | 8M/5F | 0% (0/2) |
| VV | 45 | 69±8 | 1.6±1.0 | 22M/23F | 62% (10/16) |
| **all** | **67** | **72±9** | **2.1±1.4** | **37M/30F** | **53% (10/19)** |

### Exome sequencing

We performed exome sequencing to test the hypothesis that protein-coding variants in one or more genes other than *PRNP* may cause VPSPr (Figure 2). To account for batch effects, we filtered using the lower of two allele frequencies: the filtering allele frequency (FAF) in gnomAD v4.1, which is based on the continental population with the highest frequency [27], or the allele frequency among 18,371 internal controls, which are Mendelian disease cases and unaffected family members also sequenced by the Broad Institute Genomics Platform. Implementing our allele frequency filtering strategy in the genomic browsing software *seqr* [31] resulted in no hits with >1 allele among our 67 VPSPr cases. To exclude that this null result was due to an overly aggressive filtering strategy, we also plotted the allele count in VPSPr cases against the combined filtering allele frequency obtained from our two sources (Figure 2A). This confirmed that relaxing the allele frequency filter, even by 100-fold, would not have resulted in any hits. A handful of low-frequency (AF > 0.1%) frequency variants appeared more common in VPSPr cases than controls, but all were indels, suggesting some artifact, possibly due to the brain origin of our DNA, versus the blood origin of most comparators. This analysis argues that no single protein-coding variant could cause VPSPr.

**Figure 2.**
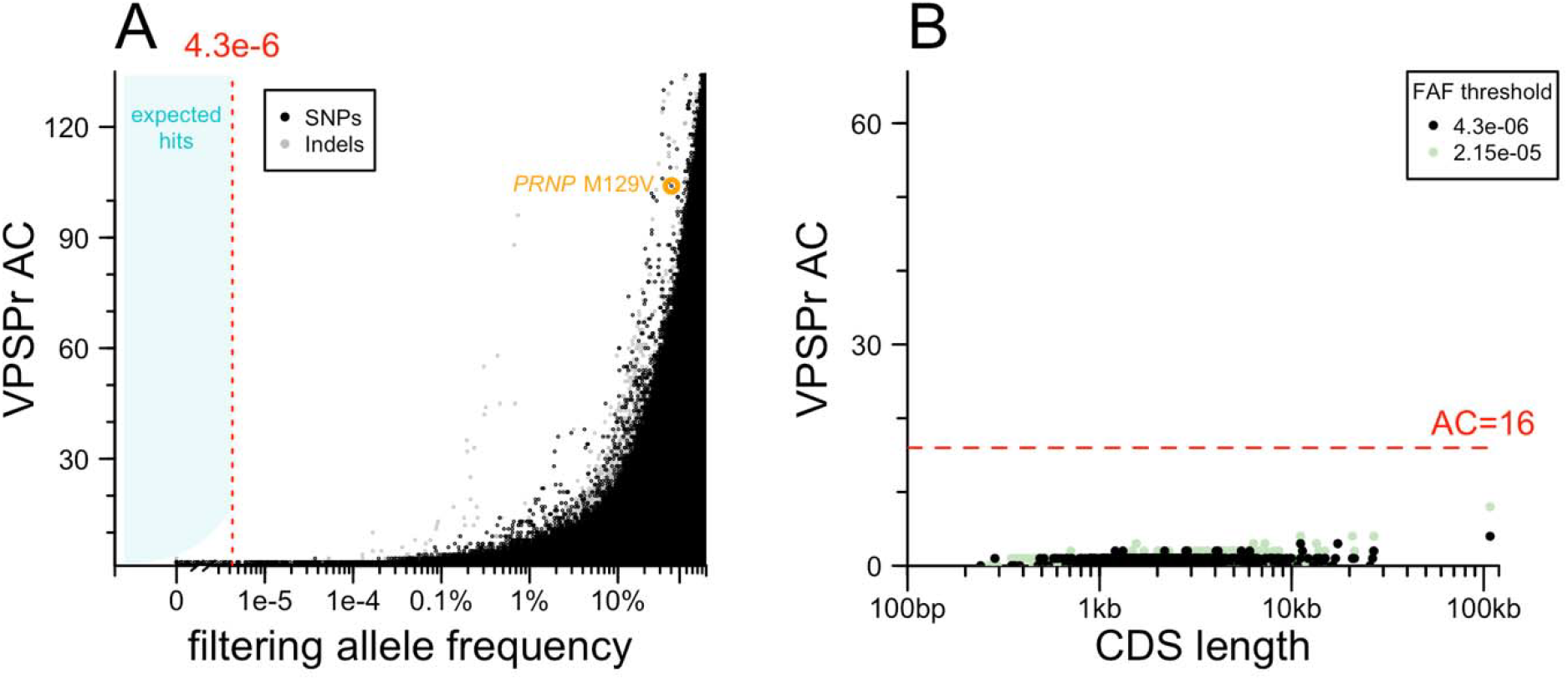
Exome sequencing results. **A)** Analysis of single variants. Each point is one variant. Allele count (AC) in VPSPr cases (y axis) versus filtering allele frequency combined from gnomAD v4.1 and internal controls for SNPs (black) and indels (gray). **B)** Analysis collapsing by gene. Each point is one gene. Total allele count (AC) of variants below frequency threshold (see inset key).

We next considered the possibility that VPSPr is genetically homogeneous but highly allelically heterogeneous — with many different causal variants, all in the same gene. Although our lack of concurrently sequenced controls does not permit a gene burden test per se, it is common to group variants by gene after applying an allele frequency filter to see if any genes are enriched for rare variants in cases. In such an approach, long genes such as *TTN* tend to be among the top hits simply because they have more opportunities for rare variation to arise. To mitigate this, we plotted the count of protein-altering variants below the filtering threshold in our VPSPr cases versus the length of coding sequence in each gene (Figure 2B). No genes had an anomalously large number of variants relative to their size. Moreover, no genes reached an allele count (AC) of 16, which is the AC at which our hypothesized OR of 1,000 intersects the filtering allele frequency (Figure 1B). Relaxing the frequency filter by 5-fold did not change this. This argues that VPSPr is not caused by protein-altering variants in any single gene.

Finally, because tau, amyloid beta, and alpha synuclein co-pathology have been observed in VPSPr brains (see Introduction), we also considered the possibility that VPSPr could be associated with familial forms of other neurodegenerative diseases. We used *seqr* [31] to search for ClinVar [32] pathogenic and likely pathogenic variants in a curated list of 25 Mendelian disease genes associated with neurodegeneration [33]. This search identified among our VPSPr cases 1 heterozygous carrier of the recessive *PRKN* R275W variant, which when homozygous causes juvenile-onset parkinsonism, as well as 1 heterozygous carrier of the *GBA* N409S variant, a common risk factor for Parkinson disease. No other hits were found, suggesting VPSPr is not associated with other familial dementias.

### Targeted sequencing of the *PRNP* locus

Prior to this study, *PRNP* coding variants had been ruled out in all of our VPSPr cases, but non-coding variation has never previously been assessed. We used targeted capture to perform deep sequencing, obtaining ~1,000X depth across almost all of a 152 kb region surrounding *PRNP*, including the lead SNPs for upstream expression quantitative trait loci (eQTLs) identified by the Genotype-Tissue Expression project (GTEx v8) [34] (Figure 3A). We compared allele counts in VPSPr to 2 control data sources: 76,215 gnomAD v4 genomes, and 298 genetic prion disease cases and carriers (see Methods). The latter were previously sequenced using the exact same targeted capture protocol, and we reasoned that they are suitable as controls because in all cases their genetic cause has already been identified as a *PRNP* coding variant, eliminating any expectation of a further non-coding contributor.

**Figure 3.**
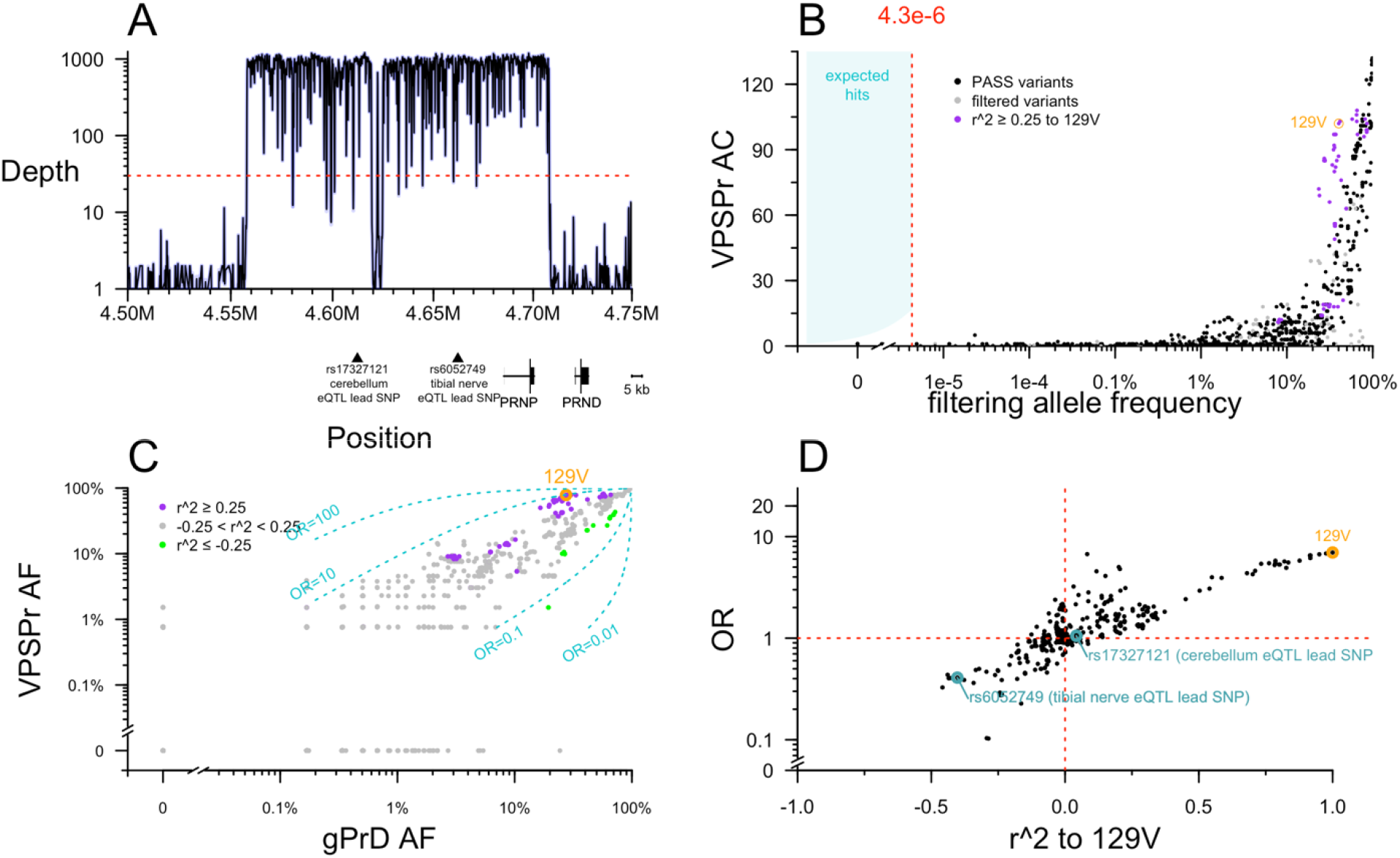
Targeted sequencing of PRNP. **A)** Targeted sequencing depth grouped by 100 base pair increments, shown relative to position of genes and relevant SNPs in GRCh38 coordinates. **B)** Analysis of single variants. Each point is one variant. Allele count (AC) in VPSPr cases (y axis) versus filtering allele frequency combined from gnomAD v4.1 genomes 76,215 and internal controls (genetic prion disease cases with known PRNP coding variants). **C)** Allele frequency (AF) among VPSPr cases (y axis) versus genetic prion disease (gPrD; x axis). Variants linked to 129V are purple and those linked to 129M are green. Dashed lines indicate ORs of 0.01, 0.1, 10, and 100. **D)** The lesser odds ratio (OR) of VPSPr versus gnomAD v4 genomes or gPrD (y axis) versus linkage to 129V (x axis).

No non-coding *PRNP* variants present in >1 VPSPr case passed the allele frequency filter. As with the exome analysis, plotting the VPSPr allele count versus control allele frequency revealed that even a dramatic relaxation of the frequency filter would not have altered this result (Figure 3B). As expected, *PRNP* M129V as well as several common variants in linkage disequilibrium with it appeared visibly enriched in VPSPr (Figure 3B).

We sought to examine further whether M129V is the sole risk variant, or whether VPSPr cases are enriched for a 129V haplotype harboring other potential risk-conferring variants. When we plotted the allele frequency of variants in and around *PRNP* in VPSPr cases versus genetic prion disease cases, most variants aligned along the diagonal representing OR = 1 (Figure 3C). Nearly all 129V-linked variants were associated with risk, with 1 < OR < 10, while nearly all 129M-linked variants were associated with protection, with 0.1 < OR < 1 (Figure 3C). To determine whether all risk associated with this haplotype could be attributed to 129V, we plotted each variant’s linkage (r^2) with 129V against its OR (Figure 3D). In this analysis, to be conservative, we used the lesser of the OR of VPSPr versus gnomAD v4 genomes or versus genetic prion disease cases. 129V had the single highest OR of any variant (OR = 7.0 relative to gnomAD non-Finnish Europeans; OR = 9.6 relative to gPrD cases), and most other variants were distributed along a diagonal where OR was proportional to r^2 (Figure 3D). The 2 upstream eQTLs fell along this diagonal (Figure 3D), suggesting that they do not have any association with VPSPr independent of linkage to 129V.

The above observations would be consistent with all risk associated to the 129V haplotype being solely attributable to 129V itself. However, we also noticed a handful of variants with anomalously high or low OR, out of proportion to their linkage to 129V (Figure 3D). The populations of our cases and controls are not perfectly matched, and we considered all ORs without regards to any particular P value threshold, so such ORs could arise by chance. To further exclude that any of these might genuinely contribute risk, we also examined several properties of all the variants in and around *PRNP* more deeply.

Plotting OR versus position did not reveal any spatially coherent cluster of high- or low-OR variants distal from M129V (Figure 4A). In sporadic CJD, codon 129 heterozygosity confers dramatic protection (OR = 0.35 in a genotypic model), but risk for 129VV is lower than 129MM, thus, in an allelic model, the 129V allele is protective (OR = 0.78) [35], opposite to VPSPr. Accordingly, all the same variants that conferred risk of VPSPr also conferred protection from sCJD according to GWAS summary statistics [35], and vice versa, all explainable by linkage to codon 129 (Figure 4B). A handful of variants exhibited OR as high as 1.3 or as low as 0.8 in the sCJD GWAS without showing any signal in VPSPr, but the converse — variants associated with VPSPr but not with sCJD — was not identified (Figure 4B), again suggesting no independent risk variants. A number of variants associated with lower risk of VPSPr are also associated with lower *PRNP* expression in tibial nerve, but the two signals were not linked by any clear positional pattern (Figure 4A and 4C). Plotting OR versus impact on tibial nerve expression aligned the variants along a diagonal, further suggesting that linkage with 129V likely underlies any appearance of an association between OR and tibial nerve expression (Figure 4D). The cerebellum eQTL is strongest further upstream of *PRNP*, in a region lacking strong OR signals (Figure 4A and 4E). 129V is not associated with cerebellar *PRNP* expression, and overall there was no correlation between cerebellum slope and OR (Figure 4F). All of the above observations are consistent with 129V being the sole *PRNP* causal risk allele for VPSPr.

**Figure 4.**
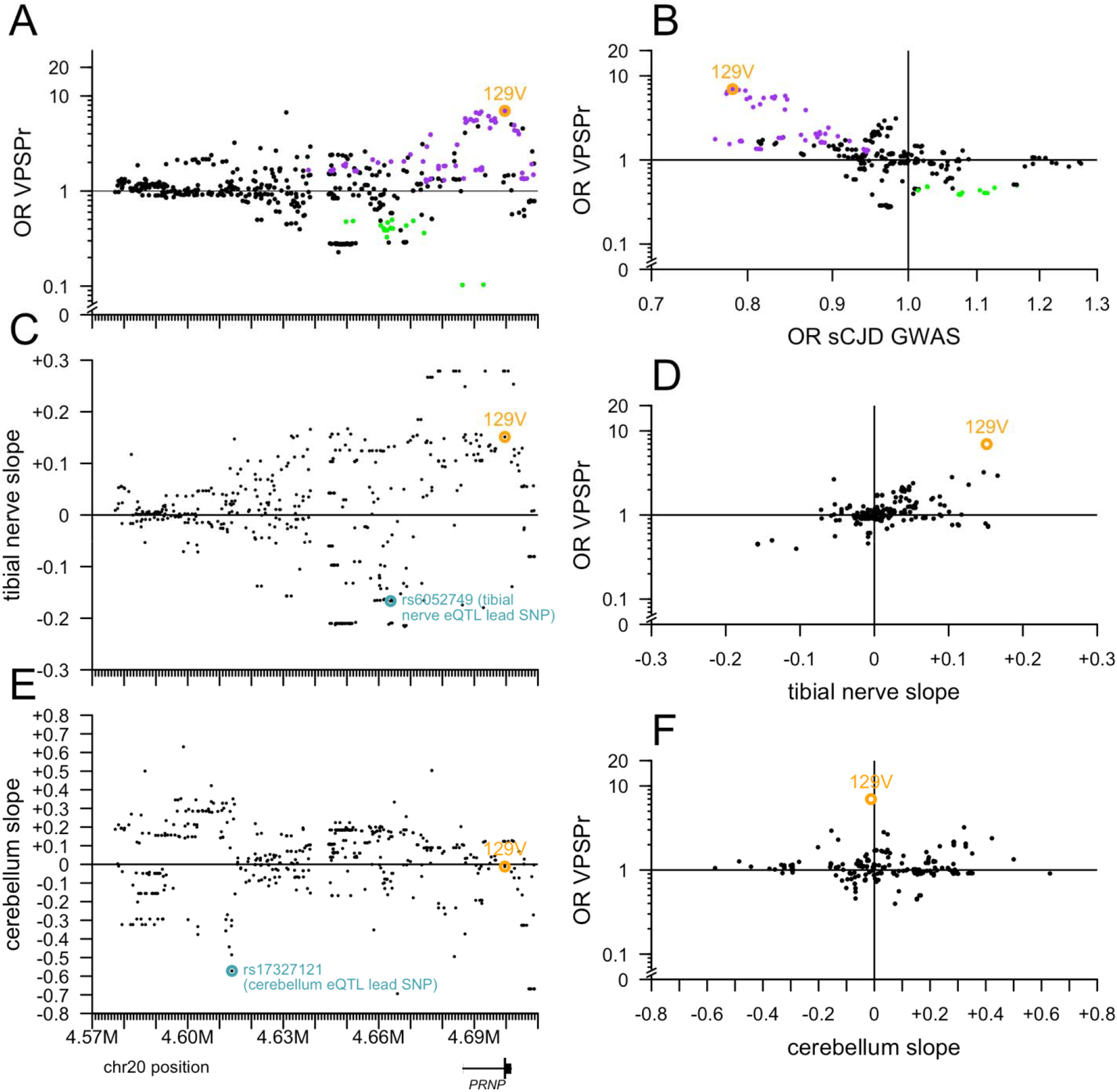
Further exploration of variants near PRNP. All targeted sequencing variants here are filtered for PASS SNPs with allele frequency >0% and <100% in both VPSPr and gPrD. **A)** The lesser odds ratio (OR) of VPSPr compared to gnomAD v4 genomes or to gPrD (y axis), versus chromosomal position (GRCh38; x axis shared with B and C). **B)** Slope of tibial nerve PRNP cis-eQTL association (GTEx v8; y axis) versus chromosomal position (x axis). **C)** Slope of cerebellum PRNP cis-eQTL association (GTEx v8; y axis) versus chromosomal position (x axis). **D)** VPSPr OR as in (A) versus the reported OR from sCJD GWAS [35]. **E)** VPSPr OR as in (A) versus tibial nerve slope as in (B). **F)** VPSPr OR as in (A) versus cerebellum slope as in (C).

While we lack genome-wide SNP data to compute a polygenic risk score or properly control for population stratification, we also checked whether lead SNPs for the top 2 hit loci in the sCJD GWAS were obviously over- or under-represented in VPSPr (Table 3). The lead SNP for *GAL3ST1* exhibited a nearly identical allele frequency (32.1%, 95%CI: 24.8-40.4%) in VPSPr as in the controls from the sCJD GWAS. For *STX6*, the alternate allele (G), which is protective (G) was found at an even lower frequency in VPSPr (50.8%, 95%CI: 42.4-59.0%) than in sCJD cases. In each case, however, confidence intervals for VPSPr allele frequency overlapped the frequencies in both sCJD cases and controls; our limited sample size does not permit determination of whether these alleles modify VPSPr risk.

**Table 3.**
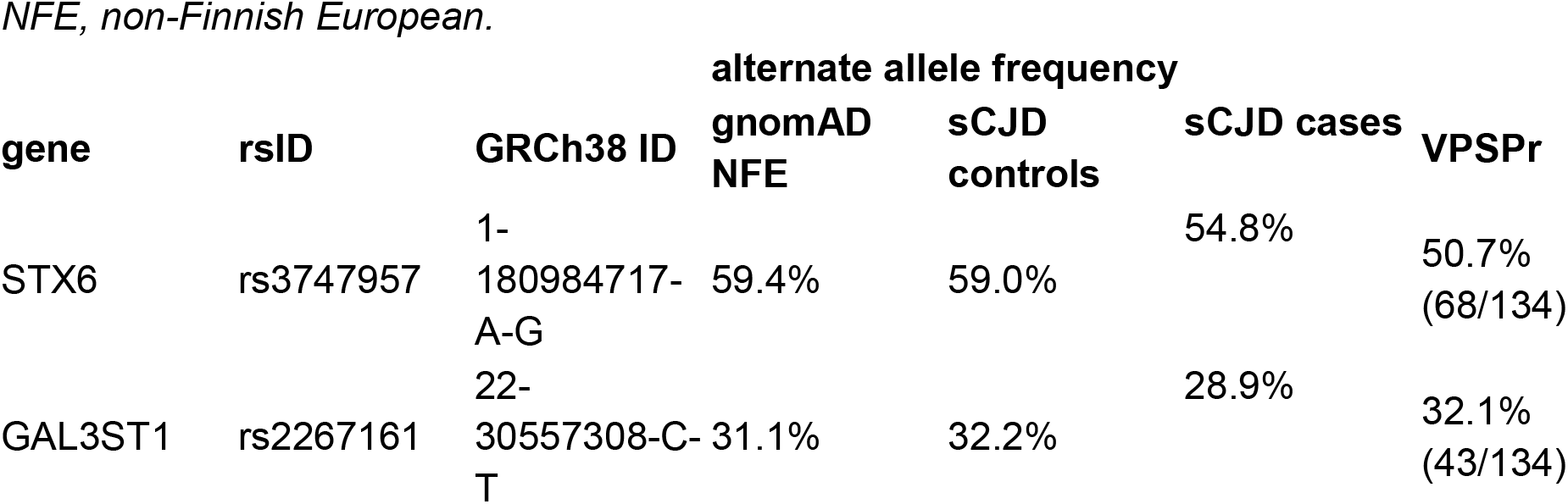
Frequencies of sCJD GWAS hit lead SNPs in VPSPr and comparator datasets. NFE, non-Finnish European.

## Discussion

Well-controlled studies have never examined the frequency of positive family history of dementia in VPSPr versus sporadic CJD or other rapidly progressive dementias. Nonetheless, the positive family history reported in 42% of VPSPr cases has led several investigators to speculate whether VPSPr has a genetic cause. Here, we examined the most obvious places where such a genetic cause might be found: protein-coding sequences of non-*PRNP* genes, and non-coding sequence in and around *PRNP*, particularly variants linked to the known risk allele 129V. We did not identify any potentially causal variants. Our data are consistent with 129V as the sole genetic risk factor for VPSPr. Note that while its OR of 7.0 would be considered as a “strong” risk factor by the standards of common disease genetics, this is not strong for a rare disease. Given that prion disease has a baseline prevalence of 0.016%, variants would need to have OR > 1,000 to cause appreciable penetrance [36].

Our study has limitations. We did not perform whole genome sequencing to look for non-coding variants in non-*PRNP* genes, and many classes of genetic variation would evade our search altogether, such as short tandem repeats (STRs) and structural variants (SVs) [37]. We chose not to sequence matched controls, so we are not able to apply the rigorous statistical controls to look for, and rule out, variants of small effect. Even near *PRNP*, our targeted sequencing does not capture all distal variation that might potentially be associated with expression of this gene.

Our data may suggest that VPSPr is a truly sporadic prion disease, with no strong genetic risk factor. If true, this would mean that the relatives of VPSPr patients are not at any inherited disease risk. The prevalence of positive family history in VPSPr remains to be explained. Given the small sample size available, it could be a chance occurrence. sCJD was also reported to have a higher rate of family history than found in matched controls [24], even though a GWAS has since shown that sCJD has relatively low heritability (h2 = 0.24 to 0.26) [35]. Alternatively, it may relate to the longer disease duration and older age of onset for VPSPr, compared to sCJD. VPSPr presentations more similar to frontotemporal dementia (FTD) or Alzheimer disease may prompt clinicians to take more complete family histories, or may prompt family members to recall family history that is actually attributable to these more common dementias.

Overall, our study supports the classification of VPSPr as a sporadic prion disease.

## Methods

### Ethical approvals

This study was approved by the Broad Institute’s Office of Research Subjects Protection (NHSR-5256).

### VPSPr samples

All VPSPr patients were autopsy-confirmed at the U.S. National Prion Disease Pathology Surveillance Center (NPDPSC, Cleveland, OH). Written informed consent was obtained from legal next of kin. DNA was extracted from frozen brain tissue.

### Control datasets

Internal controls for the exome analysis were 18,371 candidate Mendelian disease cases sequenced through the Broad Center for Mendelian Genomics (CMG). Internal controls for the targeted analysis were 257 deceased, autopsy-confirmed gPrD cases examined at NPDPSC plus 41 mostly asymptomatic participants in a gPrD mutation carrier cohort study at Mass General Hospital [38,39]; all of these individuals harbored rare *PRNP* coding variants. Population control information was taken from gnomAD v4.1 [30], comprising a total of 807,192 exomes + genomes used for the VPSPr exome analysis, including 76,215 whole genomes relevant for non-coding portions of the *PRNP* targeted sequencing analysis. Linkage disequilibrium (r^2) information was queried from gnomAD v2 using Hail [40]. eQTL information (GTEx v8) [34] was downloaded from gtexportal.org.

### Filtering allele frequency calculations

We used the approach previously described [27]. For each variant, the filtering allele frequency is the highest lower bound of the 95% confidence interval of the AF in any continental population. To convert our assumptions (Table 2) into a maximum credible allele frequency, we used the webapp available at https://cardiodb.org/allelefrequencyapp/

### Mendelian disease genes list

We used a previously curated list [33] of 25 Mendelian disease genes associated with Alzheimer’s, Parkinson’s, frontotemporal dementia/amyotrophic lateral sclerosis, and Huntington’s disease, ultimately sourced from Online Mendelian Inheritance in Man (OMIM) https://omim.org Because our question was whether familial forms of these neurodegenerative disorders are associated with risk of VPSPr, we did not include genes associated with these disorders through GWAS.

### Short read sequencing

All sequencing was performed at the Genomics Platform at the Broad Institute. Exome sequencing used the standard germline exome v6 (SGEv6) product, with 40 ng of input DNA subjected to capture by custom baits (Twist Biosciences) against 37 Mb of protein-coding exons and sequenced on NovaSeq 6000 S4 sequencers (Illumina). Targeted sequencing used a set of custom baits (Twist Biosciences) used previously [38,39], targeting a 152 kb region surrounding *PRNP*, and sequenced on NovaSeq SP (llumina). Variant calling was performed using DRAGEN 3.7.8. A resulting joint-called VCF file was imported directly into *seqr* for browser-based analysis, or parsed into a flat table format using a custom Python script to enable custom analyses in R.

### Exome analyses

Genotypes from exomes were filtered for depth (DP) ≥10 and genotype quality (GQ) ≥20. Sites were annotated using Variant Effect Predictor [41]. In cases of multiple functional annotations per variant, only the most severe consequence was retained. For grouped analyses, variants assigned to a non-missing gene symbol were filtered with MODERATE or HIGH impact, meaning missense or missense-like, and predicted protein-truncating.

### Source code, and data availability

All figures and statistics in this manuscript were generated using custom scripts in R 4.4.1. Genomic and phenotypic data for the Broad CMG cohort used for comparison is available via dbGaP accession numbers phs003047 and phs001272. Access is managed by a data access committee designated by dbGaP and is based on intended use of the requester and allowed use of the data submitter as defined by consent codes. Raw data and source code sufficient to reproduce the figures and statistics in this manuscript will be made available at https://github.com/ericminikel/vpspr

## Financial disclosure statement

This study was funded by the National Institutes of Health (R03 NS123786 to EVM). Broad CMG control cohort sequencing was funded by NIH grants (UM1 HG008900 to AODL and U01 HG011755 to AODL and MET). The *seqr* analysis platform was funded by R01 HG009141. The funders had no role in study design, data collection and analysis, decision to publish, or preparation of the manuscript.

## Acknowledgments

We thank Stephanie DiTroia for assistance with the *seqr* search strategy and Alicia Pham for assistance with dbGaP data deposition.

## Competing interests

EVM acknowledges speaking fees from Abbvie, Eli Lilly, Vertex, and Voyager; consulting fees from Alnylam and Deerfield; research support from Eli Lilly, Gate Bio, Ionis, and Sangamo. SMV acknowledges speaking fees from Abbvie, Biogen, Eli Lilly, Illumina, Ultragenyx, and Voyager; consulting fees from Alnylam and Invitae; research support from Eli Lilly, Gate Bio, Ionis, and Sangamo. BSA acknowledges research funding from CDC, NIH, CJD Foundation, and Ionis and consulting fees from Ionis, Sangamo, and Gate Bio, and royalties from Wolter Klower. AODL acknowledges consulting fees from Tome Biosciences, Ono Pharma USA, Addition Therapeutics, and Congenica; and research funding from Pacific Biosciences.

## Author contributions

Conceptualization: AODL, MET, EVM

Data Curation: YL, KK, SH, EVM

Formal Analysis: YL, EVM

Funding Acquisition: AODL, EVM

Investigation: BSA, EVM

Methodology: AODL, MET, EVM

Project Administration: BSA, SMV, EVM

Resources: KK, BSA

Software: YL, EVM

Supervision: SMV, BSA, EVM

Validation: EVM

Visualization: YL, EVM

Writing – Original Draft Preparation: EVM

Writing – Review & Editing: all authors

## Supporting Information file legends

Supplementary Figures: Surveillance Western blots of VPSPr brain material. exome_summary_by_gene.tsv.gz

- symbol: HGNC gene symbol
- cds_length: base pairs of protein-coding sequence (from gnomAD v4.1 constraint metrics)
- sum_vpspr_ac_strict: sum allele count in VPSPr cases using strict filtering frequency threshold (4.3e-6)
- sum_vpspr_ac_loose: sum allele count in VPSPr cases using loose filtering frequency threshold (2.15e-5)

exome_summary_stats.tsv.gz

- pos_id: unique GRCh38 variant ID. chromosome - 9-digit position - ref allele - alt allele
- chrom: chromosome
- pos: GRCh38 position
- ref: reference allele
- alt: alternate allele
- qual: PHRED quality score
- filter: filter status
- faf95_gnomad: 95%CI filtering allele frequency from gnomAD
- faf95_gnomad_ancestry: ancestry group contributing the filtering allele frequency
- incon_faf95: 95%CI filtering allele frequency from internal controls
- vpspr_ac: allele count in VPSPr cases
- vpspr_an: allele number in VPSPr cases
- af_pseudo: allele frequency for plotting purposes (AF=0 is plotted as AF=1e-6 to enable log scale plotting)

targeted_summary_statistics.tsv.gz

- pos_id_orig: unique GRCh38 variant ID. chromosome - 9-digit position - ref allele - alt allele, as directly pulled from multi-allelic VCF; variants are not left-aligned and minimally represented
- pos_id: unique GRCh38 variant ID. chromosome - 9-digit position - ref allele - alt allele. Variants are left-aligned and minimally represented.
- position: GRCh37 chromosomal position
- ref: reference allele
- alt: alternate allele
- qual: PHRED quality score
- filter: filter status
- faf_gprd: 95%CI filtering allele frequency from genetic prion disease (gPrD) cohort.
- group_max_faf_group: ancestry group contributing the gnomAD filtering allele frequency
- group_max_faf_frequency: 95%CI filtering allele frequency from gnomAD
- faf_combined: the greater of faf_gprd or group_max_faf_frequency
- or_gprd: the odds ratio of VPSPr compared to gPrD
- fisher_p_gprd: the Fisher exact test P value of VPSPr compared to gPrD
- ac_VPSPr: allele count in VPSPr cases
- an_VPSPr: allele number in VPSPr cases
- ac_gPrD: allele count in gPrD cohort
- an_gPrD: allele number in gPrD cohort
- p_bonf_gprd: Bonferroni-corrected version of fisher_p_gprd
- or_gprd_pseudo: or_gprd clipped to a range of [-20, 20]
- or_nfe: the odds ratio of VPSPr compared to gnomAD non-Finnish Europeans (NFE).
- fisher_p_nfe: the Fisher exact test P value of VPSPr compared to gnomAD NFE.
- ac_nfe: allele count in gnomad NFE.
- an_nfe: allele number in gnomad NFE.
- p_bonf_nfe: Bonferroni-corrected version of fisher_p_nfe
- or_nfe_pseudo: or_nfe clipped to a range of [-20, 20]
- af_scjd_gwas: allele frequency in the sCJD GWAS summary statistics (Jones 2020)
- or_scjd_gwas: odds ratio in the sCJD GWAS summary statistics (Jones 2020)
- pval_scjd_gwas: P value in the sCJD GWAS summary statistics (Jones 2020)
- r_squared_4699605: linkage disequilibrium r^2 to rs1799990 (M129V)
- r_squared_4692886: linkage disequilibrium r^2 to rs35643278 (an intronic variant showing non-Bonferroni-significant association to VPSPr in the exploratory analysis)
- slope_tibial_nerve: GTEx slope for association to tibial nerve *PRNP* expression.
- pval_nominal_tibial_nerve: GTEx nominal P value for association to tibial nerve *PRNP* expression.
- slope_cerebellum: GTEx slope for association to cerebellum *PRNP* expression.
- pval_nominal_cerebellum: GTEx nominal P value for association to cerebellum *PRNP* expression.

## Notes

### Competing Interest Statement

EVM acknowledges speaking fees from Abbvie, Eli Lilly, and Vertex; consulting fees from
Alnylam and Deerfield; research support from Eli Lilly, Gate Bio, Ionis, and Sangamo. SMV
acknowledges speaking fees from Abbvie, Biogen, Eli Lilly, Illumina, and Ultragenyx; consulting
fees from Alnylam and Invitae; research support from Eli Lilly, Gate Bio, Ionis, and Sangamo.
BSA acknowledges research funding from CDC, NIH, CJD Foundation, and Ionis and consulting
fees from Ionis, Sangamo, and Gate Bio, and royalties from Wolter Klower. AODL
acknowledges consulting fees from Tome Biosciences, Ono Pharma USA, Addition
Therapeutics, and Congenica; and research funding from Pacific Biosciences.

### Funding Statement

This study was funded by the National Institutes of Health (R03 NS123786). Broad CMG control cohort sequencing was funded by NIH grants UM1 HG008900 and U01 HG011755, and seqr analysis platform by R01 HG009141.

### Author Declarations

This study was approved by the Broad Institute Office of Research Subjects Protection (NHSR-5256)

### Summary of Updates

Response to reviewers round 1

