## Supplement for "Search for a genetic cause of variably protease-sensitive prionopathy"

Lian et al 2025

**Supplementary Figures: Surveillance Western blots of VPSPr brain material.** Per a reviewer's request, we are providing images of Western blots performed for surveillance purposes at the National Prion Disease Pathology Surveillance Center on N=10 of the VPSPr cases described in this manuscript. Diagnosis of these cases was made through a combination of Western blot, histology, and immunohistochemistry. Each blot represents 1 VPSPr case. Key: +: proteinase K-treated; -: not proteinase K-treated; F: frontal cortex (from VPSPr case); C: cerebellum (from VPSPr case); O: occipital cortex (from VPSPr case); Cx1: cortex 1 (from VPSPr case); Cx2: cortex 2 (from VPSPr Case); N: negative control (non-prion brain); T1: sCJD type 1 positive control; T2: sCJD Type 2 positive control; PTA: indicates phosphotungstic acid precipitation of $PrP^{Sc}$ was used. All blots used the 1E4 primary antibody as indicated at upper left. The volume of sample loaded is indicated at lower left. VPSPr cases generally have partially protease-resistant $PrP^{Sc}$ observed (in the "+" condition) for at least some brain regions, but usually with higher amounts of sample loaded than are required to observe $PrP^{Sc}$ in sCJD positive controls.

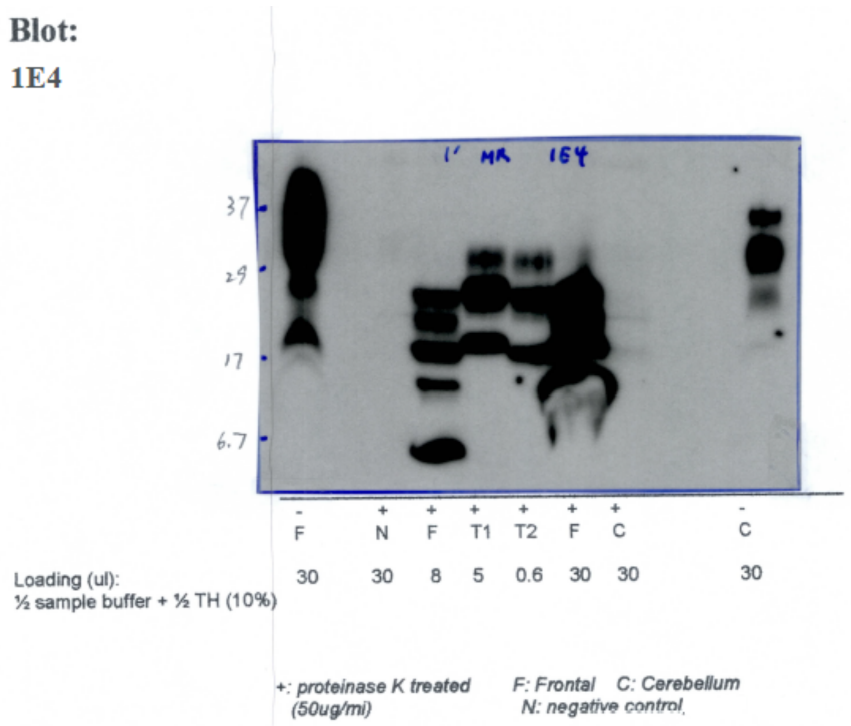

**Blot:**
**1E4**

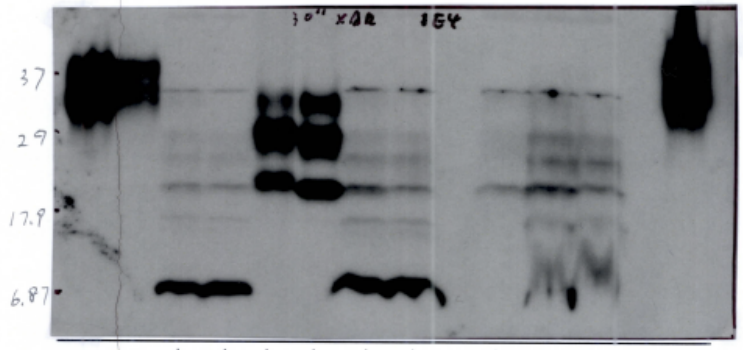


| | | | | | | | | | | | | |
|---|---|---|---|---|---|---|---|---|---|---|---|---|
| | - | - | + | + | + | + | + | + | + | + | + | - |
| | F | O | F | O | T1 | T2 | F | O | C | F | O | C |
| Loading (ul): ½ sample buffer + ½ TH (10%) | 8 | 8 | 8 | 8 | 4 | 0.4 | 15 | 15 | 30 | 30 | 30 | 30 |

*+: proteinase K treated (50ug/ml)* *F: Frontal* *O: Occipital* *C: Cerebellum*

**Blot:**
**1E4**

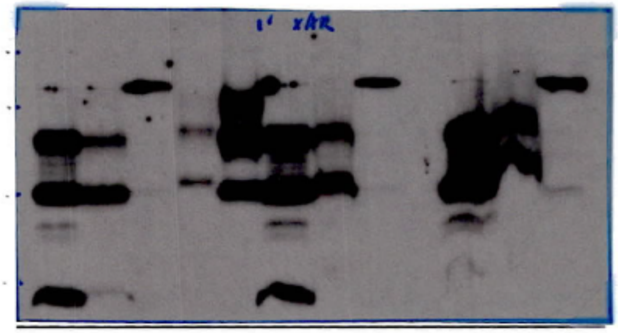


| | | | | | | | | | | | |
|---|---|---|---|---|---|---|---|---|---|---|---|
| | + | + | + | + | + | + | + | + | + | + | + |
| | F | O | C | T1 | T2 | F | O | C | F | O | C |
| Loading (ul): ½ sample buffer + ½ TH (10%) | 8 | 8 | 8 | 0.5 | 0.3 | 15 | 15 | 15 | 30 | 30 | 30 |

*+: proteinase K treated (50ug/ml)* *F: Frontal* *O: Occipital* *C: Cerebellum* *N: negative control,*

**Blot:**

**1E4**

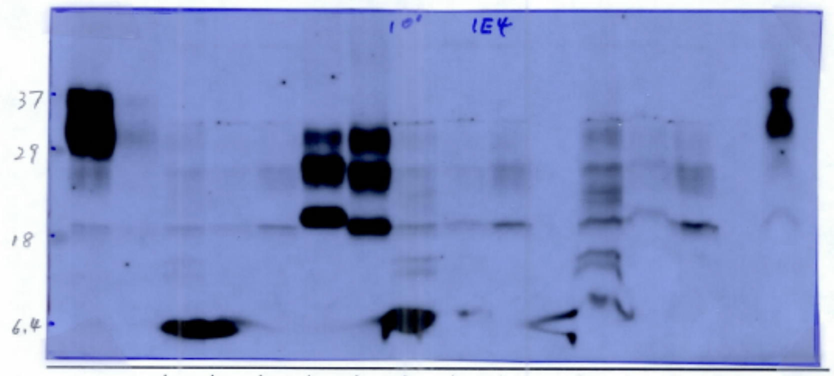


| | | | | | | | | | | | | | | |
|---|---|---|---|---|---|---|---|---|---|---|---|---|---|---|
| | - | - | + | + | + | + | + | + | + | + | + | + | + | - |
| | F | O | F | O | C | T1 | T2 | F | O | C | F | O | C | C |
| Loading (ul): ½ sample buffer + ½ TH (10%) | 8 | 8 | 8 | 8 | 8 | 4.5 | 1.2 | 15 | 15 | 15 | 30 | 30 | 30 | 30 |

*+: proteinase K treated (50ug/ml)* *F: Frontal* *O: Occipital* *C: Cerebellum*

**Blot:**
**1E4**

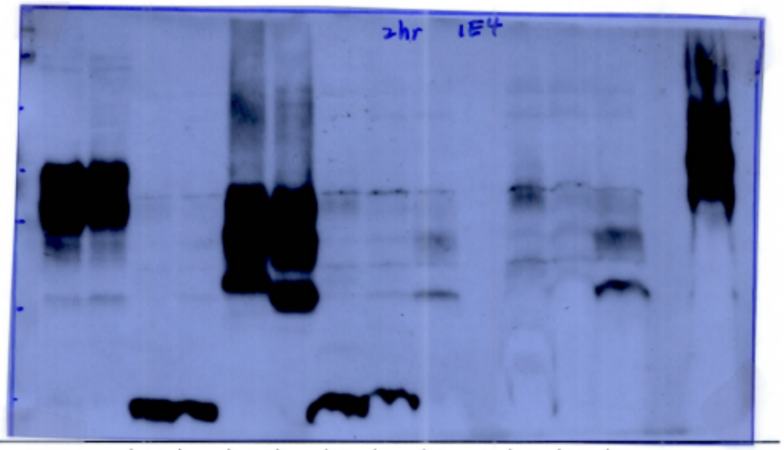


| | | | | | | | | | | | | |
|---|---|---|---|---|---|---|---|---|---|---|---|---|
| - | - | + | + | + | + | + | + | + | + | + | + | - |
| F | O | F | O | T1 | T2 | F | O | C | F | O | C | C |
| 8 | 8 | 8 | 8 | 12 | 0.7 | 15 | 15 | 15 | 30 | 30 | 30 | 30 |

Loading (ul):
½ sample buffer + ½ TH (10%)

*+: proteinase K treated (100ug/ml)*   *F: Frontal*   *O: Occipital*   *C: Cerebellum*

**Blot:**
**1E4**

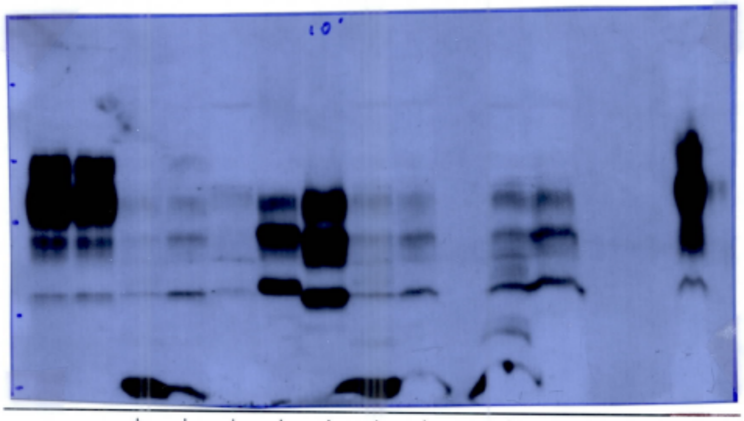


| | | | | | | | | | | | |
|---|---|---|---|---|---|---|---|---|---|---|---|
| - | - | + | + | + | + | + | + | + | + | + | - |
| F | O | F | O | C | T1 | T2 | F | O | F | O | C |
| 8 | 8 | 8 | 8 | 30 | 4.5 | 1.2 | 15 | 15 | 30 | 30 | 30 |

Loading (ul):
½ sample buffer + ½ TH (10%)

*+: proteinase K treated (50ug/ml)*   *F: Frontal*   *O: Occipital*   *C: Cerebellum*

**Blot:**
**1E4 PTA**

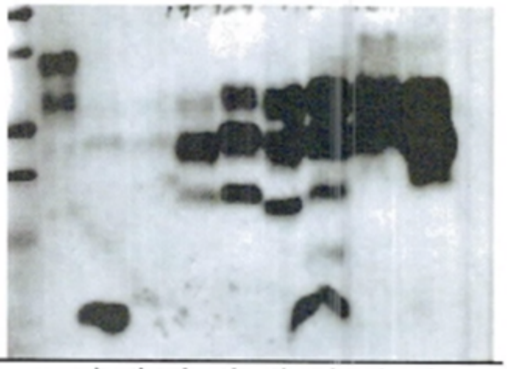

| | | | | | | | | | |
|---|---|---|---|---|---|---|---|---|---|
| | - | + | + | + | + | + | + | + | + |
| | F | F | O | C | T1 | T2 | F | O | C |
| Loading (ul): | 10 | 15 | 15 | 15 | 3.5 | 0.4 | 30 | 30 | 30 |

*+: proteinase K treated*    *F: Frontal*   *O: Occipital*   *C: Cerebellum*

**Blot:**
**1E4**

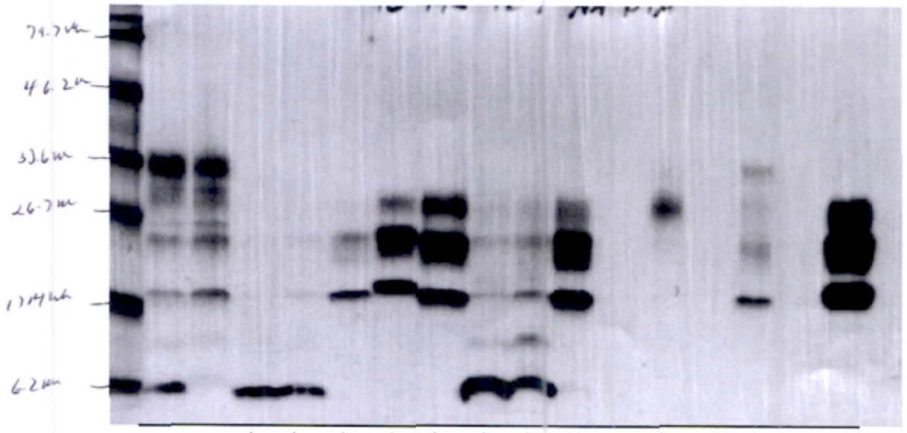

| | | | | | | | | | | | | | |
|---|---|---|---|---|---|---|---|---|---|---|---|---|---|
| | - | - | + | + | + | + | + | + | + | + | + | - | + |
| | F | O | F | O | C | T1 | T2 | F | O | C | Neg | C | Pos |
| Loading (ul):<br>½ sample buffer + ½ TH (10%) | 15 | 15 | 6 | 6 | 6 | 8 | 1 | 30 | 30 | 30 | 30 | 15 | 6 |

*+: proteinase K treated*    *F: Frontal*    *O: Occipital*   *C: Cerebellum*

**Blot:**
**1E4**

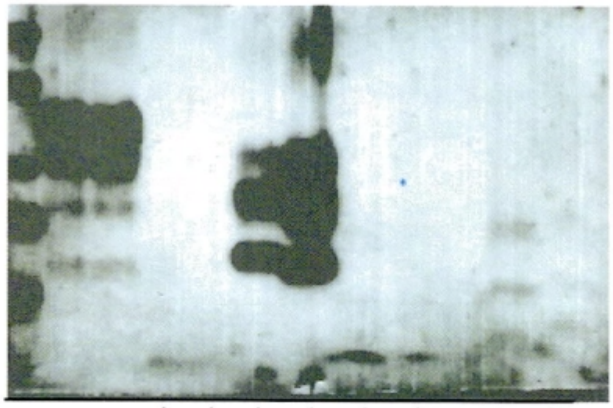

| | | | | | | | | | | |
|---|---|---|---|---|---|---|---|---|---|---|
| | - | - | + | + | + | + | + | + | + | + |
| | Cx1 | Cx2 | Cx1 | Cx2 | T1 | T2 | Cx1 | Cx2 | Cx1 | Cx2 |
| Loading (ul): ½ sample buffer + ½ TH (10%) | 8 | 8 | 2 | 2 | 2 | 0.7 | 8 | 8 | 30 | 30 |

*+: proteinase K treated*   *Cx1: Cortex1*   *Cx2: Cortex2*

**Blot:**
**1E4 PTA**

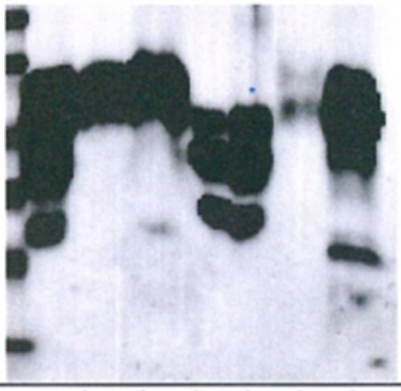

| | | | | | | | |
|---|---|---|---|---|---|---|---|
| | - | + | + | + | + | + | + |
| | O | C | O | T1 | T2 | NC | F |
| Loading (ul): | 1 | 30 | 30 | 1 | 0.2 | 30 | 25 |

*+: proteinase K treated*   *F: Frontal*   *O: Occipital*   *C: Cerebellum*
*NC: Negative control*
